# Assessing the Influence of a TikTok Aging Filter on Young Adults’ Intentions to Use Sunscreen for Skin Cancer Prevention

**DOI:** 10.1101/2025.06.13.25329616

**Authors:** David Perez, Alec McIntosh, Rui Zhang, Olga Rodriguez, Jaeil Ahn

## Abstract

**Background:** Skin cancer is among the most common cancers globally. With the rise of social media, platforms like TikTok are emerging as potential tools in public health. This study explored whether a TikTok aging filter could increase sunscreen use intentions.

**Methods:** Participants (N=230) were randomly assigned to either the experimental group (N=114), exposed to the TikTok aging filter and an educational video about ultraviolet radiation, or the control group (N=116), exposed only to the ultraviolet radiation video. A generalized linear mixed effects model was used to assess the relationship between group assignment, demographic variables, and attitudes toward sunscreen use.

**Results:** The median participant age was 22 years; 56% were female, 56% were non-White, and 92% were non-Hispanic. Multivariable analyses showed significant increases in sunscreen use intentions in both groups (OR: 6.17, p<0.001), with the experimental group showing a greater increase (OR: 4.26, p<0.001). Awareness of sunscreen benefits (OR: 4.36, p < 0.001) and concern about aging from sun exposure (OR: 6.50, p < 0.001) also increased, with no significant differences between groups.

**Conclusion:** Interactive and user-tailored visualizations can more effectively enhance sunscreen use intentions than video formats alone.

## INTRODUCTION

Skin cancer remains one of the most frequently diagnosed cancers in the world, accounting for one in every three cancer diagnoses.^1^ In the United States, the incidence and prevalence rates of melanoma and non-melanoma skin cancers have increased since 1990.^2^ Ultraviolet radiation (UVR) exposure, primarily from the sun, constitutes the biggest risk factor for the development of skin cancers, including melanoma, basal cell carcinoma, and squamous cell carcinoma.^3^ Protective behaviors against UVR, such as the use of sunscreen, remain an effective and modifiable behavior for reducing the risk of developing skin cancer.^4^

Engaging in UVR protective behaviors is especially important during young adulthood, as sunburn and UVR exposure tend to peak around the age of twenty.^5^ With the growing use of social media among this demographic, especially young adults, platforms like TikTok have arisen as potential tools for encouraging sun protection.^6^ Previous research has shown that exposure to certain types of content on TikTok can influence psychological developments such as body dissatisfaction and the internalization of beauty ideals.^7,8^ These findings highlight how influential visual content on TikTok can shape attitudes and behaviors.^7,8^ This influence, while often associated with negative outcomes like disordered eating behaviors and body image concerns, also suggests that TikTok can be leveraged for positive behavior change.

Recent studies emphasize the underexplored potential of TikTok as a channel for public health communication.^9^ The global popularity of the platform along with the attention that its trends garner make it particularly promising in encouraging sunscreen use among young adults.^10^ Among these trends is a Time Travel Filter (TTF) that simulates the user’s progressive aging process across multiple decades. By generating visual outcomes that may be hastened by unprotected sun exposure, it not only exploits the increased cosmetic concern generated by digital media but also presents a strategic avenue for public health efforts on UVR protection.^11^

The aim of this study was to evaluate whether the use of a TTF could enhance intentions to use sunscreen among young adults, focusing on the prevention of premature aging to indirectly reduce skin cancer risk.

## METHODS

This study employed a quasi-experimental design and was conducted at a private university in Washington, D.C. We recruited 230 students aged 18-34 who were enrolled at the university. Research assistants (RA), trained specifically for this study, assessed participant eligibility and managed enrollment at various on-campus locations. To be eligible, participants had to meet the following criteria: (1) be 18 years of age or older, and (2) be currently enrolled at the university. Participants were enrolled from May 24, 2024 to August 22, 2024. Enrollment concluded upon obtaining our desired sample size (N=230). This study received exemption by the Georgetown University Institutional Review Board (IRB ID: STUDY00007686). Verbal consent was obtained from all participants and witnessed by the enrolling RA before participation.

The primary objective of the study was to evaluate the effect of a TTF on attitudes and intentions towards sunscreen use. Secondary objectives included understanding participants’ baseline sun protection knowledge and practices, and investigating the relationship between TikTok usage and intended behaviors.

To address these objectives, participants completed a pre-survey that gathered demographic information, sun exposure practices, knowledge of sunscreen benefits and their concern about the effects of UVR exposure on aging (**Supp Table 1**). Participants subsequently watched a 43- second educational TikTok video regarding the effects of solar radiation on aging and skin cancer risk, with (experimental) or without (control) exposure to a TikTok TTF. Participants then completed a post-survey about their understanding and intentions regarding sun exposure. To link pre- and post-surveys, participants were given a participant identification number based on the research assistant’s initials and the participant’s order (e.g., DP’s third participant used ’DP03’ as their participant identification on both the pre- and post-surveys). The control group was composed of participants with an odd identification number and the experimental group consisted of participants with an even identification number.

**Table 1:** Study cohort characteristics by experiment groups.

| **Variable** | **Total**<br>**N=230** | **Experimental**<br>**N=114** | **Control**<br>**N=116** | **P-value** |
| --- | --- | --- | --- | --- |
| **Age<sup>1</sup>** | 22 (20-24) | 22 (20-25) | 22 (20-24) | 0.62 |
| **Sex** |  |  |  |  |
| Female | 129 (56%) | 64 (28%) | 65 (28%) | 0.34 |
| Male | 98 (43%) | 50 (22%) | 48 (21%) |  |
| Non-Binary | 3 (1.3%) |  | 3 (1.3%) |  |
| **Race** |  |  |  |  |
| White | 101 (44%) | 50 (22%) | 51 (22%) | 1.00 |
| Non-White | 129 (56%) | 64 (28%) | 65 (28%) |  |
| **Ethnicity** |  |  |  |  |
| Hispanic | 18 (7.8%) | 6 (2.6%) | 12 (5.2%) | 0.22 |
| Non-Hispanic | 212 (92%) | 108 (47%) | 104 (45%) |  |
| **Education Background** |  |  |  |  |
| Health related | 194 (84%) | 93 (40%) | 101 (44%) | 0.28 |
| Non-Health related | 36 (16%) | 21 (9.1%) | 15 (6.5%) |  |
| **TikTok Use** |  |  |  |  |
| Never | 97 (42%) | 42 (18%) | 55 (24%) | 0.55 |
| Once a week | 14 (6.1%) | 8 (3.5%) | 6 (2.6%) |  |
| Every few days | 17 (7.4%) | 10 (4.3%) | 7 (3.0%) |  |
| Once per day | 23 (10%) | 13 (5.7%) | 10 (4.3%) |  |
| Multiple times a day | 79 (34%) | 41 (18%) | 38 (17%) |  |

<sup>1</sup>Median (IQR); N(%)

**Table 1: Study cohort characteristics by experiment groups**
| Variable | Total<br>N=230 | Experimental<br>N=114 | Control<br>N=116 | P-value |
| --- | --- | --- | --- | --- |
| **Age<sup>1</sup>** | 22 (20-24) | 22 (20-25) | 22 (20-24) | 0.62 |
| **Sex** |  |  |  |  |
| Female | 129 (56%) | 64 (28%) | 65 (28%) | 0.34 |
| Male | 98 (43%) | 50 (22%) | 48 (21%) |  |
| Non-Binary | 3 (1.3%) |  | 3 (1.3%) |  |
| **Race** |  |  |  |  |
| White | 101 (44%) | 50 (22%) | 51 (22%) | 1.00 |
| Non-White | 129 (56%) | 64 (28%) | 65 (28%) |  |
| **Ethnicity** |  |  |  |  |
| Hispanic | 18 (7.8%) | 6 (2.6%) | 12 (5.2%) | 0.22 |
| Non-Hispanic | 212 (92%) | 108 (47%) | 104 (45%) |  |
| **Education Background** |  |  |  |  |
| Health related | 194 (84%) | 93 (40%) | 101 (44%) | 0.28 |
| Non-Health related | 36 (16%) | 21 (9.1%) | 15 (6.5%) |  |
| **TikTok Use** |  |  |  |  |
| Never | 97 (42%) | 42 (18%) | 55 (24%) | 0.55 |
| Once a week | 14 (6.1%) | 8 (3.5%) | 6 (2.6%) |  |
| Every few days | 17 (7.4%) | 10 (4.3%) | 7 (3.0%) |  |
| Once per day | 23 (10%) | 13 (5.7%) | 10 (4.3%) |  |
| Multiple times a day | 79 (34%) | 41 (18%) | 38 (17%) |  |

<sup>1</sup>Median (IQR); N(%)

The anonymized data were recorded electronically on Qualtrics. Graphical statistical analysis of the data was performed using Jamovi version 2.6.2. Descriptive statistics such as median with interquartile range (IQR) and counts with percentages were used to summarize participants’ baseline demographics and characteristics. Chi-square tests were used to determine the univariate association between race, sex, type of schools, TikTok usage, and sun protection behaviors between the control and experimental groups. For changes in attitude towards each sunscreen use survey outcome, McNemar’s tests were used for univariate analyses, respectively. For the multivariable adjusted analysis (MVA), multinomial cumulative logistic regression with random intercepts was used to assess odds ratios and their 95% confidence intervals for the intervention group, survey time, and the group × time interaction, adjusting for informative confounders selected via stepwise variable selection with p < 0.1. The cumulative logit assumption or proportional odds assumption was evaluated using likelihood ratio tests and goodness-of-fit statistics such as Akaike Information Criterion. Additional sensitivity analyses were performed using 1) subsets where participants without potential improvement at post-survey were excluded and 2) including all covariates regardless of their statistical significance. Statistical significance was determined using a two-sided p-value of < 0.05 or < 0.0125 (Bonferroni correction), as appropriate. All multivariable analyses were performed using SAS procedures *GLIMMIX* and *NLMIXED ver 9.4*.

## RESULTS

There was a median age of 22 years across all groups. Females comprised 56% of all participants, and made up the majority in both the control (28%) and experimental (28%) groups, while males accounted for 21% and 22%, respectively (**Table 1**). Non-White individuals represented 56% of the total group, with a similar distribution in both the control (28%) and experimental (28%) groups (**Table 1**). The majority of participants identified as Non-Hispanic (92%), and 84% of the total sample attended health-related schools within Georgetown University (**Table 1**). When asked about the frequency of sunbathing, 13.4% reported sunbathing at least once per month or more, while 47.4% reported sunbathing only while on vacation (**Figure 1A**). A total of 23.9% reported using sunscreen no more than once per month (**Figure 1B**). Baseline awareness of sunscreen benefits was moderate or lower among 66.5% of respondents (**Figure 1C**). Half of the participants expressed slight concern or no concern about the effects of aging from sun exposure (**Figure 1D**).

**Figure 1.**
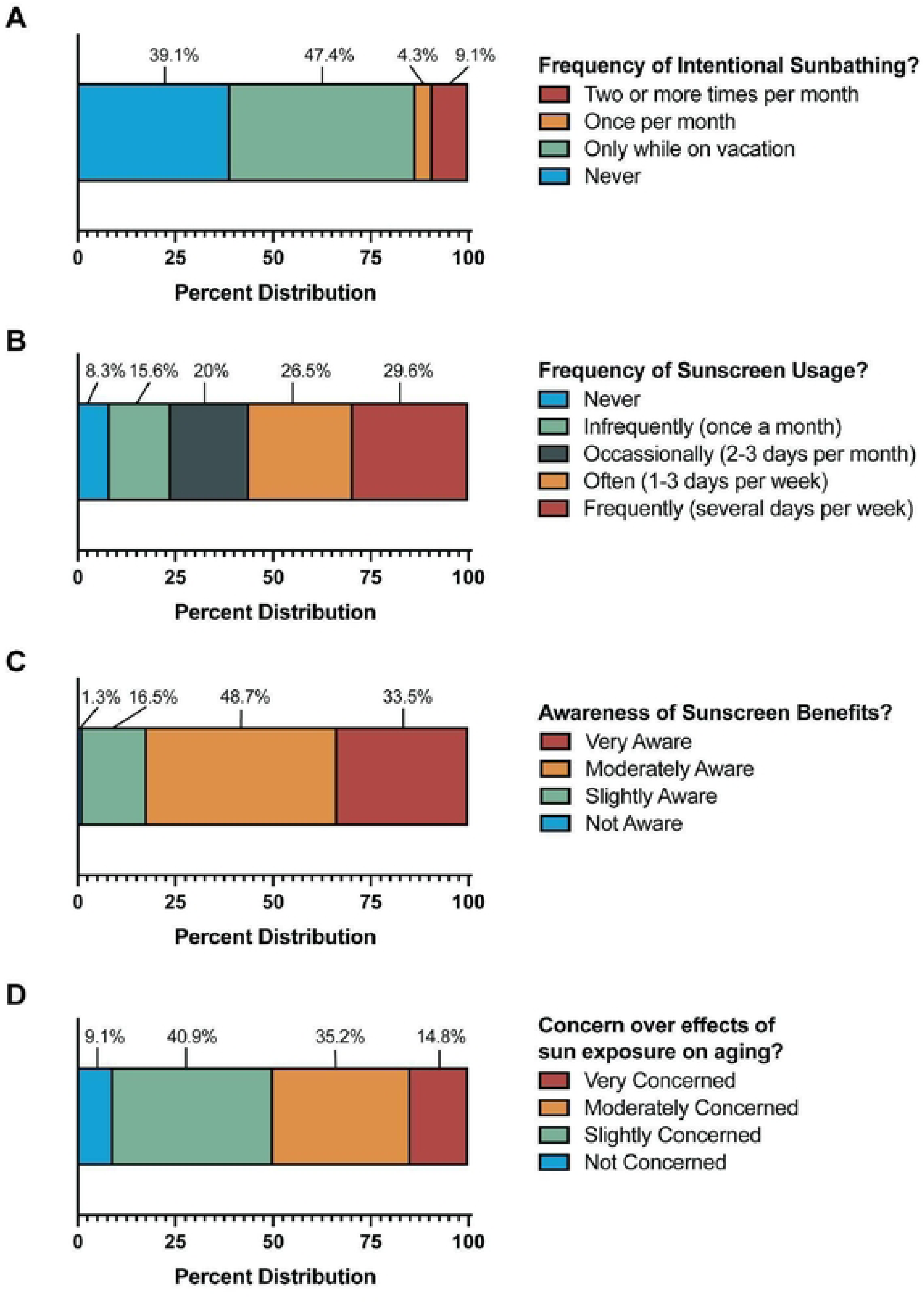
Stacked bar graphs depicting the distribution of responses to pre-intervention survey questions pertaining to baseline (**A**) Sunbathing behavior, (**B**) Sunscreen usage, (**C**) Awareness of sunscreen benefits, and (**D**) Concerns over the effects of sun exposure on aging. Percentage of individual answers of all 230 participants are displayed.

Following the intervention, the MVA showed that among the experimental group, there was no significant change between participants’ self-reported sunbathing frequency in the pre-survey and their intended sunbathing frequency reported in the post-survey. (p=0.46) (**Figure 2A**). Independent of group assignment, race emerged as a significant predictor as White individuals were significantly more likely to report an intention to sunbathe compared to Non-White individuals (OR: 4.75, 95% CI: 2.40–9.37, p<0.001) (**Figure 2A**). In contrast to sunbathing behavior, intentions to use sunscreen significantly increased in both groups following the intervention (OR: 6.17, 95% CI: 3.56–10.7, p<0.001), with a more pronounced increase in the experimental group (OR: 4.26, 95% CI: 1.87–9.74, p<0.001) (**Figure 2B**). Despite the overall increase in sunscreen use intentions, male participants were significantly less likely to express an intention to use sunscreen (OR: 0.10, 95% CI: 0.05–0.20, p<0.001) (**Figure 2B**). While sunscreen intentions varied by gender, awareness of sunscreen benefits significantly increased in both groups (OR: 4.36, 95% CI: 2.52–7.54, p<0.001), with no significant differences between the groups (p = 0.68) (**Figure 3A**). At the demographic level, White individuals, older students, and females were significantly more likely to be aware of sunscreen benefits (p=0.05, 0.02, 0.005, respectively) (**Figure 3A**). In addition to significant increases in sunscreen awareness, there was a significant increase in concern about the aging effects of sun exposure in both groups (OR: 6.50, 95% CI: 3.84–11.0, p<0.001), with males showing less concern compared to females (OR: 0.34, 95% CI: 0.20-0.59, p<0.001) (**Figure 3B**). Our findings were unchanged when adjusting for all covariates assessed and the subset analyses performed (**Figures S1-S4**).

**Figure 2.**
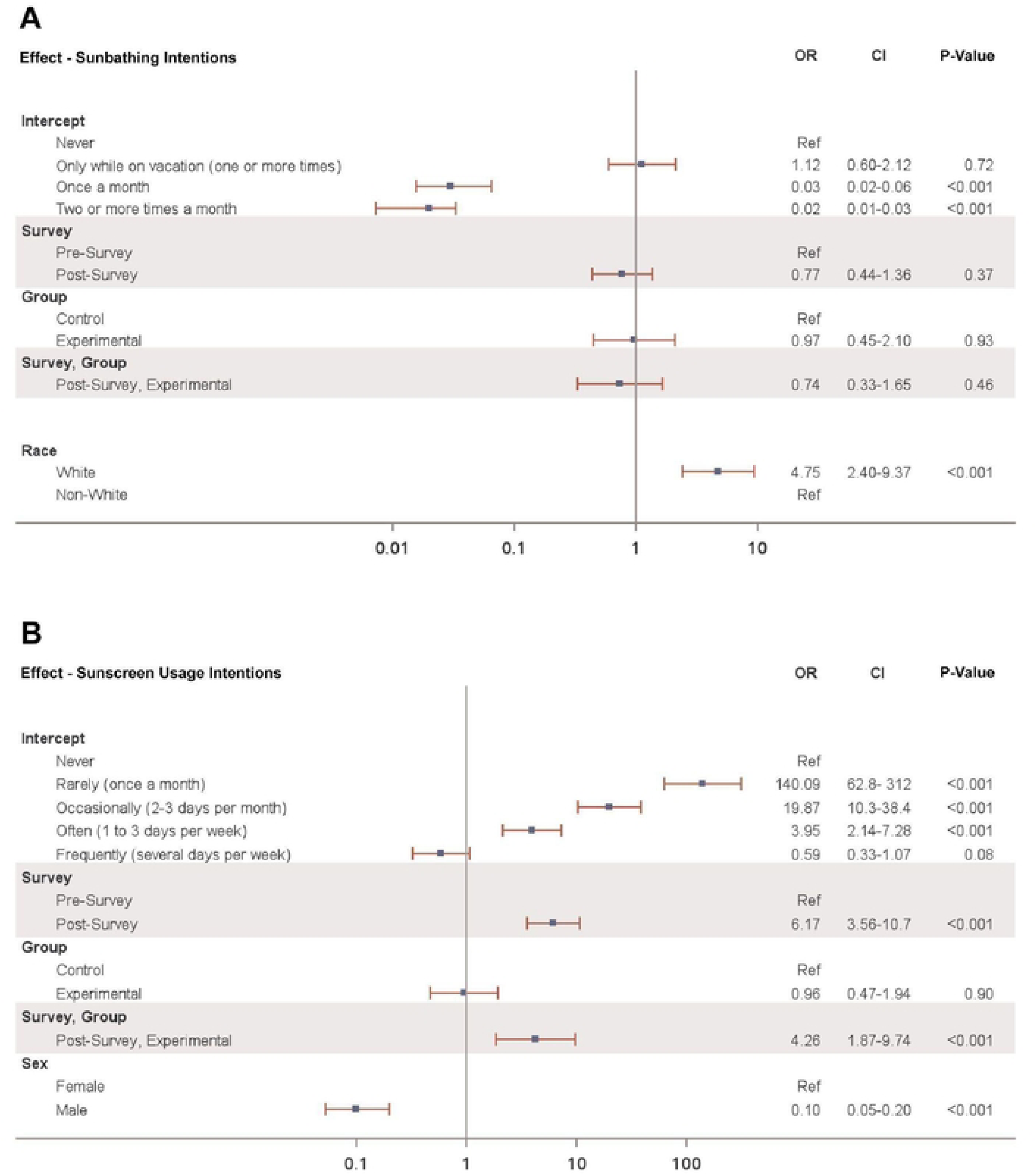
Multivariable-Adjusted Analysis reporting Odds Ratio (OR) with 95% CI for reported sun protective behaviors using only significant covariates for (**A**) Sunbathing intentions and (**B**) Sunscreen usage intentions, respectively. OR > 1 are indicative of greater sunbathing and sunscreen use intentions, respectively.

**Figure 3.**
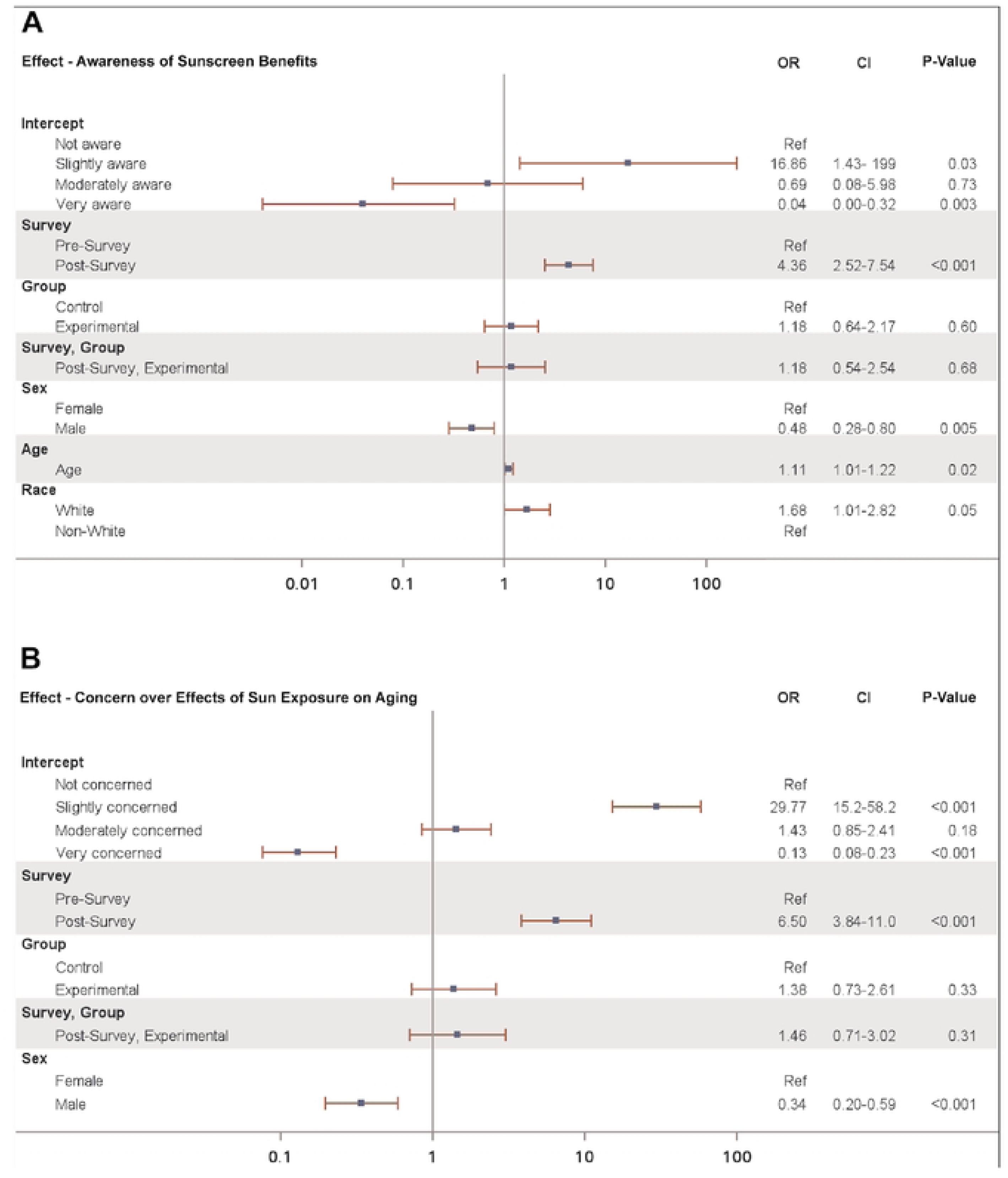
Multivariable-Adjusted Analysis reporting Odds Ratio (OR) with 95% CI for reported understanding of sun damage using only significant covariates for participant (**A**) Awareness of sunscreen benefits and participant (**B**) Concern over effects of sun exposure on aging, respectively. OR > 1 are indicative of greater awareness and concern, respectively.

## DISCUSSION

In this study, our objective was to assess whether immersive TikTok media could increase young adults’ intentions to use sunscreen. Results showed that although both groups experienced significant improvements in intended sunscreen use, the experimental group demonstrated a greater increase in sunscreen use intentions. In addition to changes in behavioral intentions, both groups reported greater awareness regarding the benefits of sunscreen and increased concern about the effects of sun exposure on skin aging. These findings suggest that while educational videos can raise awareness and promote proactive attitudes, the addition of personalized, emotionally engaging content, such as artificial intelligence (AI) aging filters, may play a more powerful role in influencing behavioral intentions. This finding aligns with previous research by McCashin et al,^9^ which emphasizes TikTok’s value as a tool for public health promotion. In recognition of the psychological risks associated with TikTok, our study focused on a positive intervention through careful framing of health-related content to encourage proactive sun protection.^7,8^

At the demographic level, the relatively young age of the participants corresponds with the critical period for sun protection behaviors cited by Thieden et al,^5^ as sunburns and excessive UVR exposure tend to peak in young adulthood. This suggests that TikTok, with its interactive features, could serve as an effective platform to influence sun protection behaviors among this age group. Non-White participants, who reported lower sunbathing rates than White participants, could benefit from interventions addressing incidental UVR exposure, while interventions for White participants could focus on the risks of frequent sunbathing and incorporate AI-based tools that help users visualize additional effects of UVR damage, such as sunburns and the development of skin cancer. Gender-specific content could also be useful, as males showed lower sunscreen use intentions. For this group, incorporating physician-featured short-video messages, which have been shown to significantly increase behavioral intentions, may be helpful in increasing sunscreen use intentions within an AI-based approach.^12^

It is important to note that regression analyses, including those with all covariates, showed that the frequency of TikTok use did not significantly predict sunbathing intentions, sunscreen use intentions, awareness of sunscreen benefits, or concern about sun exposure and aging (**Figures S1-S4**). This suggests that the intervention’s effectiveness was not confounded by participants’ prior engagement with the platform, but rather by the content itself. As such, even infrequent or non-users of TikTok may still benefit from similar interventions.

Limitations of this study include the reliance on self-reported measures, which introduces potential biases such as social desirability and recall bias. Participants may have over-reported positive behaviors, such as sunscreen application or concern about aging effects, particularly in the experimental group after the intervention. In addition, the sample was primarily composed of young students from a private university, and is not sufficiently representative of nationwide demographics, specifically age and racial groups or those from other types of institutions. Intrinsically, the findings are most applicable to similar age groups and university settings, limiting broader application. While the study shows short-term changes in sunscreen use intentions, the long-term impact of TikTok-based interventions on sustained behavior change remains uncertain. We also acknowledge that the use of unvalidated survey items limits the reliability of the data collected. Future research should explore strategies for promoting long-term engagement and measuring the durability of these behavior changes.

## CONCLUSION

Our findings highlight the potential of AI content in enhancing the impact of health education, as it may more effectively promote positive behavioral change than traditional video formats alone.

Personalized interventions like TikTok’s TTF can make abstract risks feel more immediate, thereby strengthening behavioral intentions. Demographic factors, such as gender, remain important in influencing responsiveness. Expanding and tailoring these approaches could contribute to addressing broad health challenges such as the rising incidence and prevalence rates of skin cancer.

## Data Availability

The data underlying the results presented in the study are available from David Perez/Jaeil Ahn

## ACKNOWLEDGEMENTS

We are deeply grateful to Nesreen Shahrour, Janessa M. Mendoza, Karen Sun, Christine Chow, and Woori Lee for their assistance in data collection.

---

**Supplemental Table 1.** Pre- and Post-Intervention Survey Questions.

| **Pre-Survey** |
| --- |
| 1. How old are you? |
| 2. How do you describe yourself?<br>- Male<br>- Female<br>- Non-binary<br>- Transgender<br>- Other |
| 3. What is your race?<br>- White<br>- Black or African American<br>- American Indian or Alaska Native<br>- Asian<br>- Native Hawaiian or Pacific Islander<br>- Middle Eastern<br>- Mixed race<br>- Other (please specify) |
| 4. Do you identify as Hispanic/Latinx?<br>- Yes<br>- No |
| 5. Are you enrolled in a Georgetown health school (School of Medicine, School of Nursing, School of Health, etc.)?<br>- Yes<br>- No |
| 6. How often do you currently use sunscreen?<br>- Frequently (several days per week)<br>- Often (1 to 3 days per week)<br>- Occasionally (2-3 days per month)<br>- Rarely (once a month)<br>- Never |
| 7. How often do you sunbathe (laying in the sun to tan)?<br>- Two or more times a month<br>- Once a month<br>- Only while on vacation (one or more times)<br>- Never |
| 8. How aware are you about the benefits of sunscreen?<br>- Not aware<br>- Slightly aware |
| - Moderately aware<br>- Very aware |
| 9. How concerned are you about the effects of sun exposure on aging?<br>- Not concerned<br>- Slightly concerned<br>- Moderately concerned<br>- Very concerned |
| 10. How often do you use TikTok?<br>- Multiple times a day<br>- Once per day<br>- Every few days<br>- Once a week<br>- Never |
| **Post-Survey** |
| 1. How often do you plan on sunbathing (laying in the sun to tan)?<br>- Two or more times a month<br>- Once a month<br>- Only while on vacation (one or more times)<br>- Never |
| 2. How often do you plan on using sunscreen?<br>- Frequently (several days per week)<br>- Often (1 to 3 days per week)<br>- Occasionally (2-3 days per month)<br>- Rarely (once a month)<br>- Never |
| 3. How concerned are you about the effects of sun exposure on aging?<br>- Not concerned<br>- Slightly concerned<br>- Moderately concerned<br>- Very concerned |
| 4. How aware are you about the benefits of sunscreen?<br>- Not aware<br>- Slightly aware<br>- Moderately aware<br>- Very aware |

## Notes

### Competing Interest Statement

The authors have declared no competing interest.

### Funding Statement

The author(s) received no specific funding for this work.

### Author Declarations

This study received exemption by the Georgetown University Institutional Review Board (IRB ID: STUDY00007686). Verbal consent was obtained from all participants before participation.

